# Resting state fMRI reveals pervasive thalamic hyperactivity and default mode network hypoactivity in epilepsy: Systematic review and meta-analysis

**DOI:** 10.1101/2023.08.21.23294356

**Authors:** Yang Qiao, Cong Fu, Na Zhao, Matthew Lock, Zhen Yuan, Yu-Feng Zang

## Abstract

**Objective:** This coordinate-based meta-analysis incorporates studies employing local metrics like amplitude of low frequency fluctuation (ALFF) and regional homogeneity (ReHo), and studies utilizing independent component analysis (ICA) to probe the default mode network (DMN) in epilepsy.

**Methods:** A comprehensive literature search was conducted in PubMed, Embase, and Web of Science to identify relevant studies published up to July 2022. We included all studies that compared RS-fMRI local activity in epileptic patients and healthy controls.

**Results:** From 644 identified studies, 54 were included in the meta-analyses. Our analysis revealed a consistent increase in local activity in the right mediodorsal thalamus (MDT), bilateral medial temporal lobe, and bilateral sensorimotor cortex. Conversely, a notable decrease in local activity was observed within the DMN regions, including the posterior cingulate cortex (PCC)/precuneus, medial prefrontal cortex, and bilateral inferior parietal lobule. Furthermore, a significant negative correlation between abnormal activity in the right MDT and PCC was identified through our meta-correlation analysis.

**Conclusions:** The findings from our study provide compelling evidence of increased local activity in the mediodorsal thalamus and decreased activity in the default mode network in epilepsy. These observations could potentially advance our understanding of epilepsy’s neural underpinnings and guide targeted intervention strategies.

What is already known on this topic?

Epilepsy involves abnormal brain activity, recurrent seizures, and possible thalamic and DMN involvement.

What this study adds?

This study reveals consistent thalamic hyperactivity, DMN hypoactivity in various epilepsy types, and a significant negative correlation between these activities.

How this study might affect research, practice, and/or policy?

Decreased DMN activity and increased thalamus activity could clarify epilepsy pathogenesis and suggest precise, targeted interventions.

## 1. Introduction

Epilepsy is a common chronic neurological disease with a lifetime incidence of about 8 people per thousand (1). Although various antiepileptic drugs (AEDs) have been the preferred treatment (2), drug resistance and side effects are a substantial issue (3). About half of all patients do not have good control after the first treatment, and about 35% of patients do not respond to AEDs (4). For patients with refractory epilepsy, precise positioning therapies were adapted, including surgical resection and precise localized modulation. Before surgical resection, intracranial electroencephalography (iEEG) was the widely accepted gold standard for localizing epileptic foci, however, it was challenging to determine the exact position and number of electrodes needed. Currently, deep brain stimulation (DBS) (5) and transcranial magnetic stimulation (TMS) (6,7) have been used as localized modulation techniques for the treatment of epilepsy. A key issue for all the surgical resection, iEEG, DBS, and TMS is individualized precise localization of the abnormal activity.

Resting-state functional magnetic resonance imaging (rs-fMRI) has the advantage of non-invasiveness, fairly good spatial and temporal resolution, and available in most hospitals (8,9). A few rs-fMRI analytic methods, e.g., amplitude of low frequency fluctuation (ALFF) (10) and regional homogeneity (ReHo) (11), are performed in a way of “voxel-wise whole-brain” analysis and are available for coordinate-based meta-analysis (12). Some coordinate-based meta-analyses based on these rs-fMRI local activity parameters have confirmed abnormal brain activity in a few disorders. For example, a meta-analysis found decreased ALFF in the posterior cingulate cortex (PCC) in Alzheimer’s disease and mild cognitive impairment (13,14). This same area has been reported to show increased amyloid deposition and decreased glucose metabolism in Alzheimer’s disease or mild cognitive impairment in a voxel-wise meta-analysis of positron emission tomography (PET) studies (15). Similar rs-fMRI approaches have been widely performed in epilepsy studies. There have been roughly more than 1,000 rs-fMRI papers, however, no related coordinate-based meta-analysis has been reported about abnormal local brain activity of epilepsy. We discovered only one meta-analytic paper of rs-fMRI functional connectivity (FC) studies on epilepsy, which reported decreased FC in the default mode network (DMN), a set of regions usually showing task independent deactivation, across different epilepsies (16). Moreover, studies of local activity have also reported reduced ALFF or ReHo in DMN (17–20). These convergent decreases in activity in the DMN provide strong evidence to support its neural correlates, like task-negative activation, as DMN activity in epilepsy is suspended by increased abnormal activity.

Seizures is caused by abnormal overfiring of the brain, so the abnormal elevation found on rs-fMRI should be specific for epilepsy. Some studies have found that local ALFF or ReHo increased in the seizure onset zone (SOZ). Among them, medial temporal lobe epilepsy (mTLE) lesions are relatively consistent since multiple mTLE studies have found increased brain activity of the medial temporal lobe (MTL) (21). In addition, thalamus, a key subcortical structure, plays a crucial role in regulating cortical neuronal excitability and inhibition, thereby influencing the stability of neural networks (22,23). When abnormal discharges from the SOZ propagate to the thalamus, the thalamus integrates and modulates these signals, affecting the activity of other cortical areas and subcortical structures. In this regard, the thalamus has been implicated in both the initiation and spread of seizures (24–26). An EEG-fMRI review has found that thalamus had shown increased activity in different type of epilepsy studies (27). Also, several rs-fMRI studies have found abnormally elevated ALFF or ReHo on thalamic activity (17,28). Therefore, we speculate that meta-analysis based on local activity may find abnormal increased local activity in the thalamus.

In rs-fMRI studies, independent component analysis (ICA) is usually taken as a brain network analytic method to study the spontaneous brain activity of a specific network. However, ICA studies typically report results in a voxel-wise manner in all brain regions of that specific network. Therefore, the ICA results have been combined with rs-fMRI local metrics, e.g., ReHo and ALFF in previous coordinate-based meta-analysis on brain disorders. For example, Hannawi and colleagues combined ICA of the DMN (ICA-DMN hereafter) and ALFF in a coordinate based meta-analysis and found decreased activity in the PCC in disorders of consciousness (29).

In the current meta-analysis, we collected and analyzed epilepsy studies using rs-fMRI with ALFF, ReHo, and ICA-DMN. Our hypothesis is that epilepsy has not only increased local activity in the thalamus but also decreased activity in the DMN, and furthermore, the increased and decreased activity should be highly correlated. The results of this meta-analysis could help precisely locate the pathophysiology of epilepsy, and thus help discover biomarkers of epilepsy to assist in neurosurgical planning, precise localized brain modulation therapy, drug targeting, and further our understanding of the biology of epilepsy.

## 2. Methods

### 2.1. Search strategy

The literature search was conducted following the Preferred Reporting Items for Systematic Reviews and Meta-Analyses (PRISMA) guideline (30). Relevant articles published prior to July 2022 were identified via a literature search on PubMed, Embase, and Web of Science using the following keywords:

(ALFF OR (amplitude of low frequency fluctuation OR (ReHo OR (regional homogeneity)))) AND (epilepsy OR (epilepsies OR (seizure))) AND (magnetic resonance OR (MRI))

(ICA OR (independent component analysis)) AND (DMN OR (default mode network)) AND (epilepsy OR (epilepsies OR (seizure))) AND (magnetic resonance OR (MRI))

The inclusion criteria are as follows: 1) study must be human research; 2) study must report results as precise coordinates in standardized space (Montreal Neurological Institute (MNI) or standard Talairach space); 3) study should contain resting-state fMRI and all the analyses must be based on a whole brain analysis; 4) the contrast should be between-group comparison (patients and healthy control group); 5) study must contain multiple relevant contrasts counted as independent cases. The current study was registered on Prospective Register of Systematic Reviews PROSPERO database (CRD42021276600).

### 2.2. Selection of studies

After removing duplicate papers, two authors (Y.Q. and C.F.) screened all the remaining references by title and abstracts independently. References that did not fulfill the inclusion criteria were excluded. At this stage, disagreements were well discussed and arbitrated by a third reviewer (N.Z.). The PRISMA flow chart illustrating the number of studies screened, excluded, and other selection details (Fig 1). A total of 54 published articles with ALFF, ReHo and ICA-DMN satisfied the above inclusion criteria.

**Fig 1.**
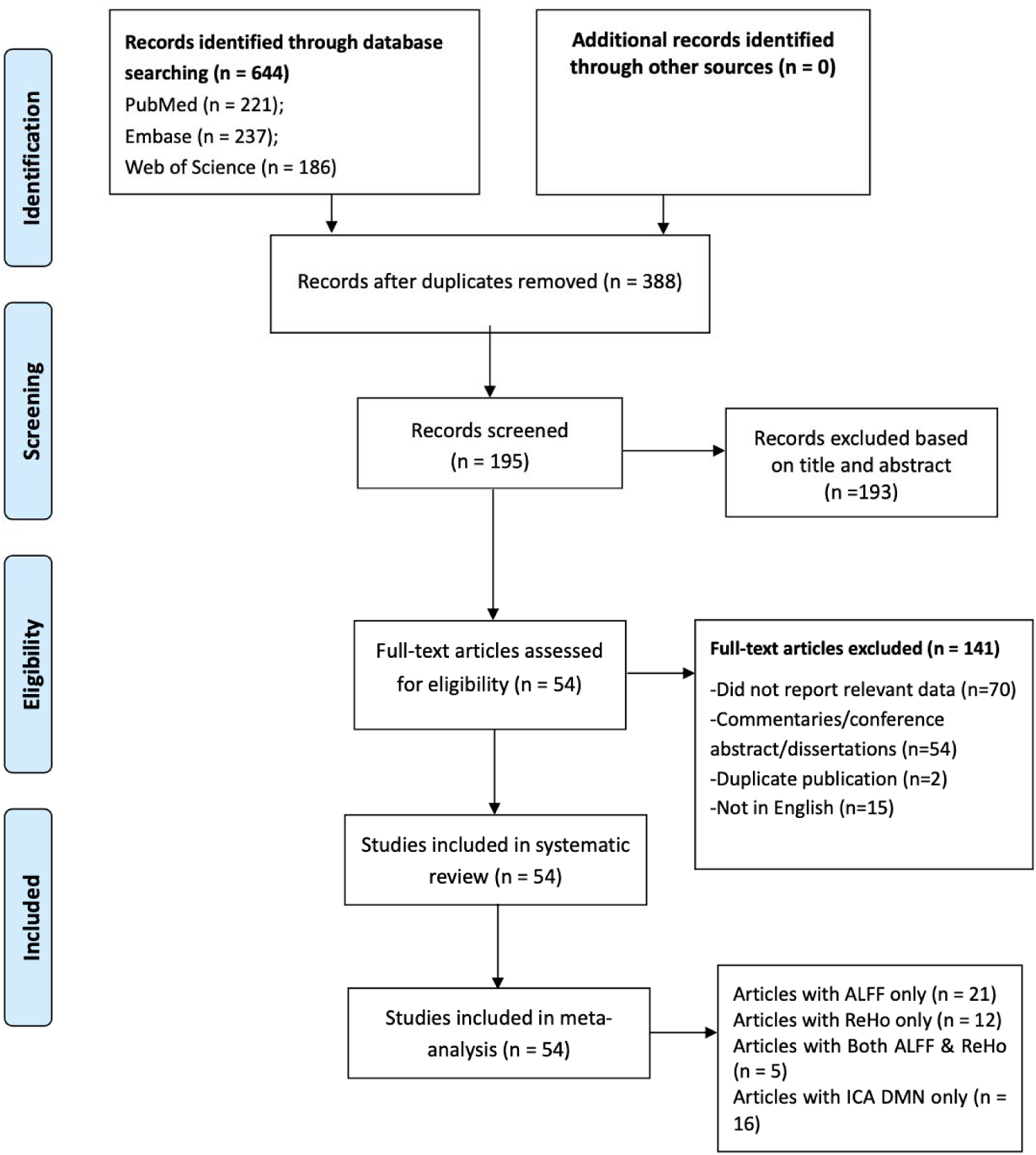
PRISMA flow chart. A total of 54 studies met the criteria for this meta-analysis, including ALFF only (n = 21), ReHo only (n = 12), both ALFF and ReHo (n =5), and ICA-DMN (n =16).

### 2.3. ALE analyses

GingerALE software (version 3.0.2) was used to carry out random effects activation likelihood estimation (ALE) meta-analyses (31) (http://brainmap.org/ale). Coordinates converting (if needed) from Talairach space to MNI space was performed by the plugin function, Convert Foci, based on Lancaster’s transformation (32).

The ALE meta is a method of coordinate based meta-analysis which aims to estimate the likelihood that a peak lies in any given voxel. This method works on multiple independent experiments with consideration of participants’ number in each experiment. The ALE meta consists of four steps. First, the ALE values are calculated for each voxel in the brain and result in an ALE map. Second, a permutation test is performed to determine the null distribution of the ALE statistic at each voxel. Third, a P value image is produced from the previous step, which can be used to set a significance threshold on the ALE map. Finally, a cluster analysis is performed on the threshold map, based on the minimum volume that is specified in the previous step. Different descriptive and summary maps of regional brain fluctuation for different groups were generated by preserving three-dimensional spatial coordinates that define the areas functioned in the original studies at the level of each voxel.

We ran four sets of analyses, first a combination of all metrics, then each metric including ALFF, ReHo, and ICA-DMN. The significance of statistical p value maps was determined using a frequently used uncorrected p<0.001 (33). For 15 studies reporting more than one relevant contrast for a specific meta-analysis (e.g., subgroup 1, subgroup 2, and healthy control), we pooled the coordinates for relevant contrasts into two different cases. For 5 studies contained both ALFF and ReHo results, we assigned them into the ALFF group and only ALFF results were extracted for ALE calculation. Also, we performed a sensitivity analysis to assess the generality of effects. Finally, we ran additional test for both TLE and non-TLE studies since TLE is the most common type of epilepsy. We believe this classifying approach can help further understand the intrinsic brain activity for both negative and positive alternation.

### 2.4 Meta-correlation

We hypothesized that there would be increased activity in the thalamus and a decreased activity in the DMN. Task fMRI studies have confirmed that the decreased activity in the DMN is correlated with the cognitive load (34). Thus, we assumed that there might be a correlation between the decreased and increased activity. We selected studies with both increased local activity in the thalamus and decreased local activity in the DMN. The *t* value and sample size were extracted to calculate Cohen’s *d* value (35). If a study reported more than one cluster in the thalamus or in the DMN, the Cohen’s *d* value was averaged inside thalamus or DMN separately. Linear correlation was performed between the Cohen’s *d* value in the thalamus with that in the DMN.

## 3. Results

### 3.1 Regions with significant abnormal local activity

We firstly combined ALFF, ReHo, and ICA-DMN coordinates and generated a general map of meta-analytic results (Fig 2, Table 1). As predicted, decreased local activity was found in the DMN regions including the PCC/precuneus, medial prefrontal cortex (mPFC), and bilateral inferior parietal lobule (IPL). Increased activity mainly located at the bilateral MTL, right thalamus, and bilateral sensorimotor cortex.

**Fig 2.**
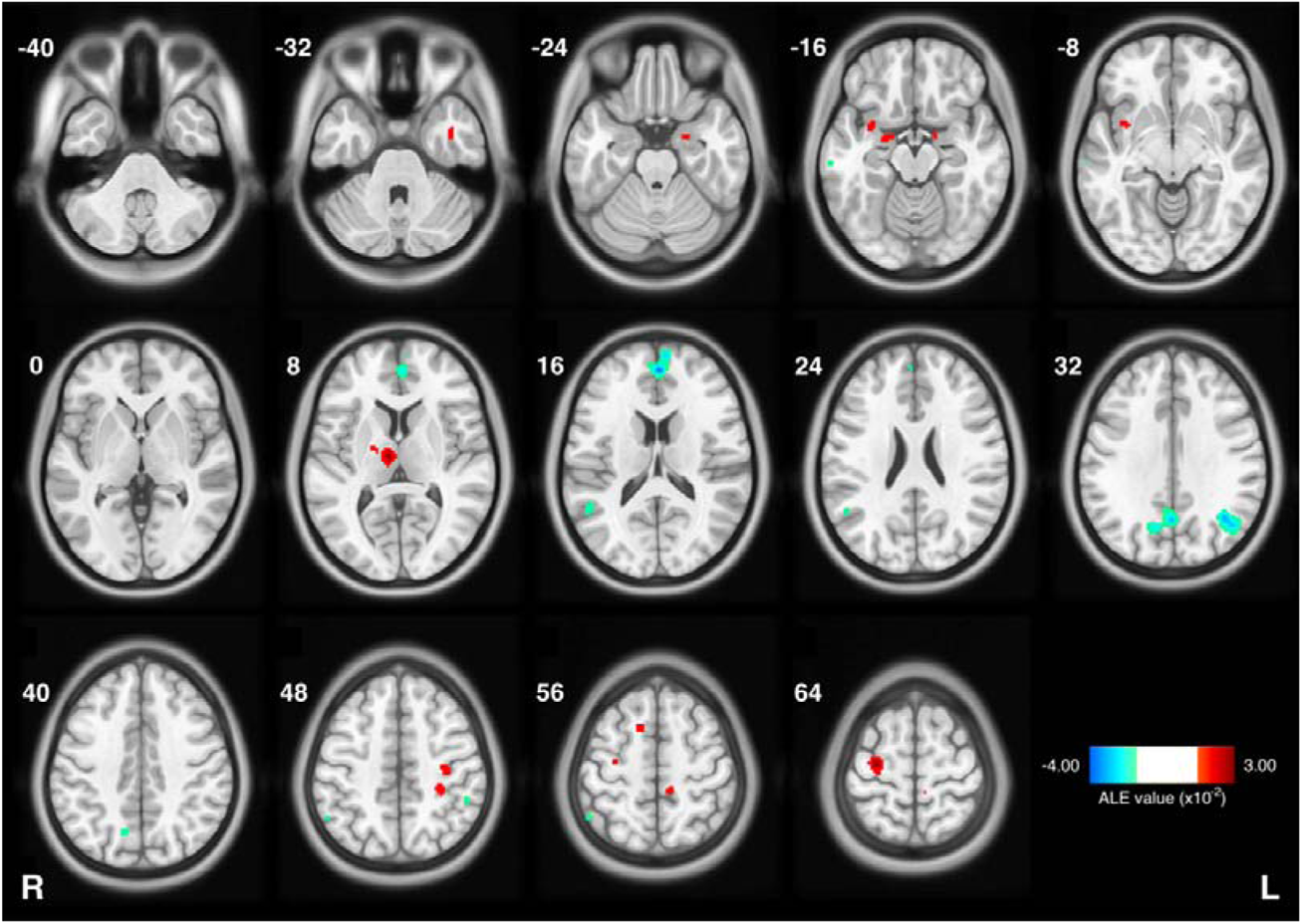
Results of meta-analysis on combined amplitude of low frequency fluctuation (ALFF) (n = 26 studies), regional homogeneity (ReHo) (n = 12 studies), and ICA-DMN (n =16 studies). For studies containing both ALFF and ReHo, we assigned them into the ALFF group and only ALFF results were extracted for activation likelihood estimation (ALE) calculation. The color bar represents ALE values, and for the decreased results, we multiply ALE value by −1 for better illustration (p<0.001 corresponding negative ALE<−1.36×10^−2^ and positive ALE>1.29×10^−2^). Red color indicates increased local activity and blue color means decreased. The numbers beside each slice represents z coordinate in Montreal Neurological Institute (MNI) space. The letters “L” and “R” means left and right separately.

**Table 1.** Brain regions showing different local activity in combined meta results (Fig 2). MNI: Montreal Neurological Institute; ALE: activation likelihood estimation.

| Cluster Size (mm<sup>3</sup>) | Side | Brain region | Peak MNI coordinates |  |  | ALE value (10<sup>-2</sup>) |
| --- | --- | --- | --- | --- | --- | --- |
|  |  |  | x | y | z |  |
| 2584 | Left | Pregenual Anterior Cingulate Cortex | -2 | 50 | 14 | 3.78 |
| 2176 | Right | Precuneus | 12 | -66 | 36 | 3.68 |
| 2160 | Left | Angular | -40 | -60 | 32 | 3.15 |
| 1240 | Right | Mediodorsal Lateral Thalamus | 8 | -12 | 8 | 2.86 |
| 1032 | Right | Precentral Gyrus | 30 | -14 | 62 | 2.98 |
| 784 | Right | Middle Temporal Gyrus | 50 | -52 | 20 | 2.77 |
| 640 | Right | Insula | 34 | 6 | -12 | 1.95 |
| 496 | Left | Postcentral Gyrus | -30 | -34 | 50 | 2.30 |
| 416 | Right | Inferior Parietal Gyrus | 52 | -56 | 52 | 2.05 |
| 344 | Left | Precuneus | -10 | -36 | 60 | 2.10 |
| 320 | Right | Hippocampus | 20 | -4 | -18 | 1.79 |
| 312 | Right | Supplementary Motor Area | 12 | 12 | 58 | 2.13 |
| 264 | Left | Postcentral Gyrus | -36 | -20 | 48 | 1.93 |
| 256 | Left | Amygdala | -22 | -2 | -22 | 1.57 |
| 224 | Left | Inferior Temporal Gyrus | -40 | 0 | -34 | 1.82 |
| 216 | Left | Inferior Parietal Gyrus | -50 | -42 | 46 | 1.89 |
| 208 | Right | Middle Temporal Gyrus | 64 | -22 | -12 | 1.67 |

We further explored the consistency across ALFF, ReHo, and ICA-DMN methods. We first performed meta-analyses of ALFF, ReHo, and ICA-DMN separately. Because ICA-DMN only contained results in the DMN, which showed decreased activity, we performed an overlap analysis of ALFF and ReHo to increase results and found an overlapping increase in local activity in the thalamus (Fig. 3A). For the DMN regions, we performed the overlapping analysis of ALFF, ReHo, and ICA-DMN. All three metrics showed decreased local activity in the DMN regions including the PCC and mPFC (Fig. 3B, Supp Fig 1).

**Fig 3.**
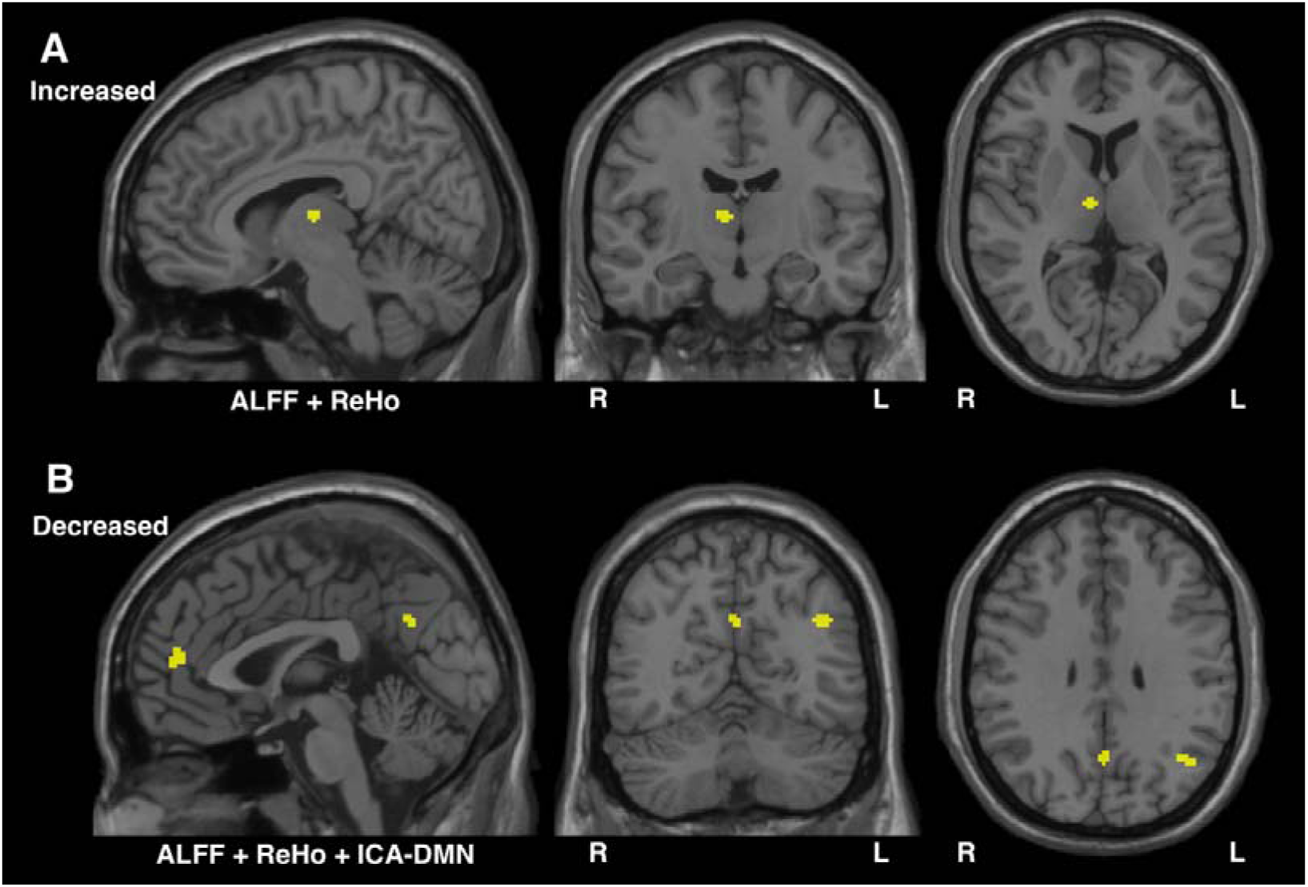
Overlapped ALFF, ReHo and ICA-DMN subgroups meta results. A: overlapped regions with increased local activity; B: overlapped regions with decreased local activity. The letters “L” and “R” means left and right respectively.

### 3.2 TLE and non-TLE comparison

Temporal lobe epilepsy (TLE), the most common type of focal epilepsy, has been widely studied with rs-fMRI (36). We have found increased activity in the medial temporal lobe without epilepsy type into account. One problem is that the increased activity in the thalamus may be limited to patients with TLE. Thus, we divided all enrolled studies into two groups, TLE group (16 studies) and non-TLE group (23 studies). We were interested in the overlapping results of the two groups.

The DMN decreased activity was overlapped on the anterior cingulate cortex. As expected, when the non-TLE patients were excluded, the TLE patients showed more abnormal increased regions in MTL, but interestingly, the non-TLE patients still had increased area in MTL. Additionally, in both types, there were abnormal increased areas in the right thalamus, which overlapped spatially. However, the thalamus showed more increased results in non-TLE group (Fig 4, SuppFig2.).

**Fig 4.**
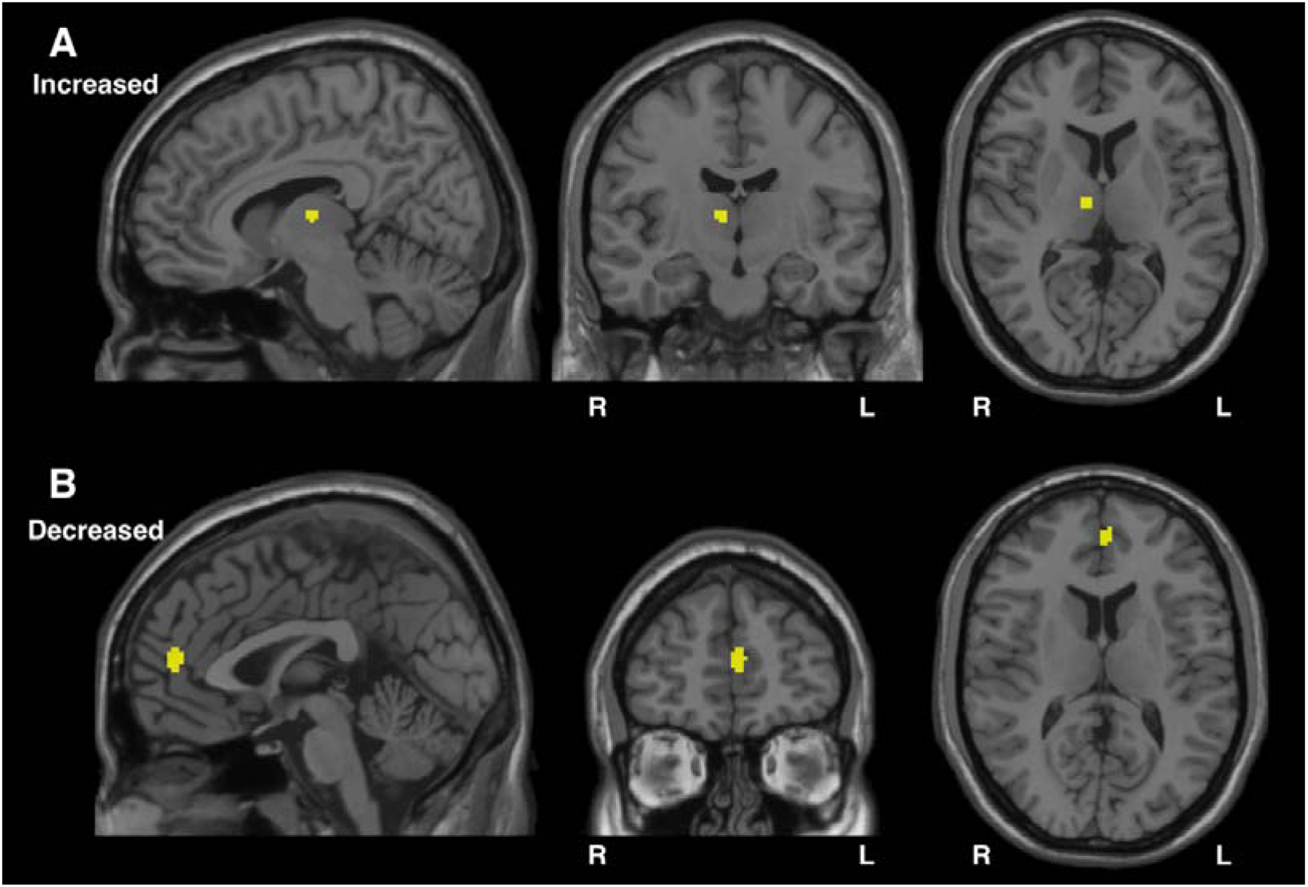
Overlapped TLE and non-TLE groups meta results. A: overlapped regions with increased local activity; B: overlapped regions with decreased local activity. The letters “L” and “R” means left and right separately.

### 3.3 Meta-correlation

A total of 8 studies (with 10 contrasts) containing both increased activity in the thalamus and decreased activity in the DMN. It should be noted that no study reported both increased or both decreased activity in the thalamus and DMN. Meta-correlation showed that the thalamic activity was negatively correlated with the DMN activity (R^2^ = 0.8121) (Fig 5).

**Fig 5.**
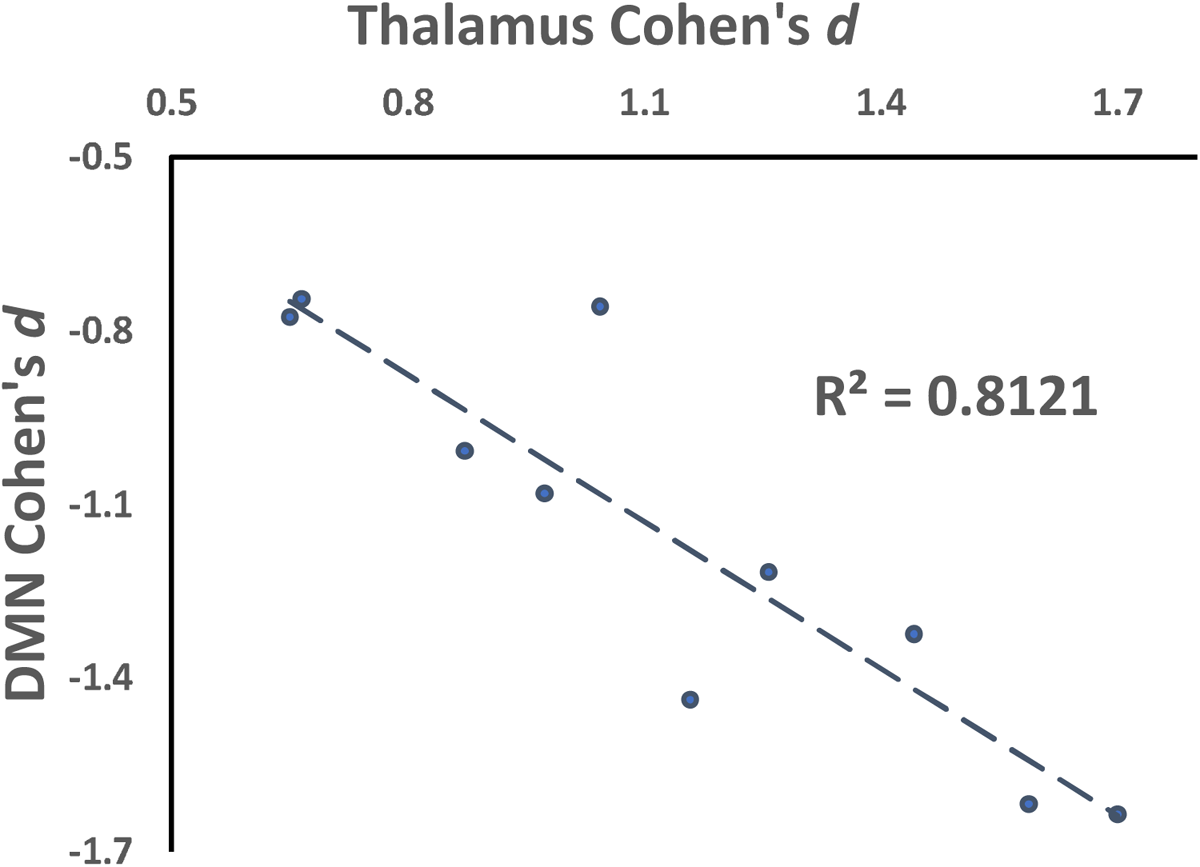
Linear correlation plot of thalamic and DMN activities. The Cohen’s *d* value was averaged inside thalamus or DMN separately.

### 3.4 Sensitivity analysis

To address potential bias of sample, we repeated the ALE analyses with the removal of large sample studies. The criterion of large sample study is number of participants (patients plus healthy controls) larger than 80, and 8 studies were removed for this analysis. The results kept consistent with those that combined all of the studies’ results (Fig 6).

**Fig 6.**
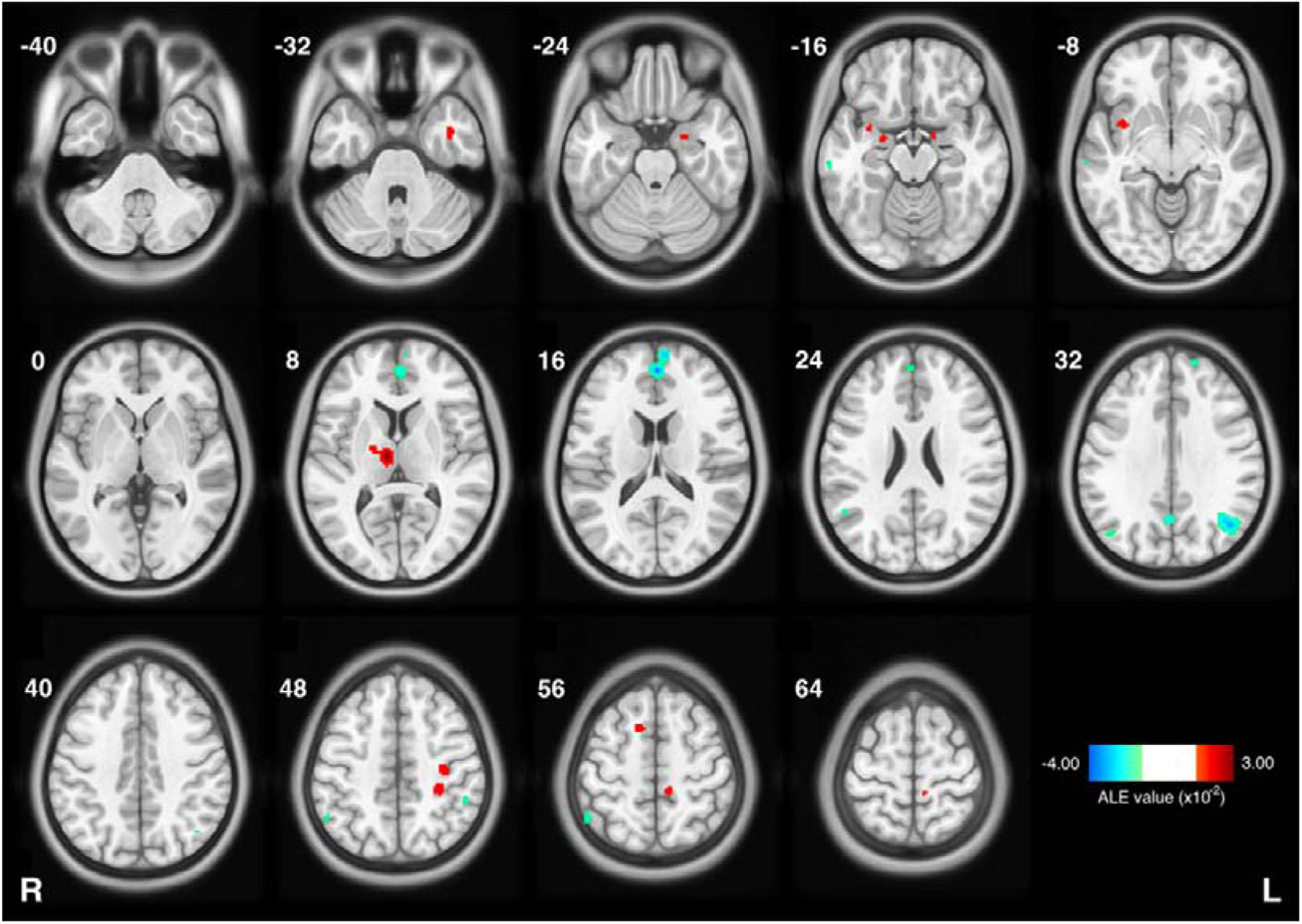
Combing ALFF, ReHo and ICA-DMN meta results with large sample studies removed. The color bar represents ALE values, and for the decreased results, we multiply ALE value by −1 for better illustration (p<0.001 corresponding negative ALE<−1.29×10^−2^ and positive ALE>1.23×10^−2^). Red color indicates increased local activity and blue color means decreased. The numbers beside each slice represents z coordinate in MNI space. The letters “L” and “R” means left and right separately.

## 4. Discussion

By conducting a systematic review and coordinate-based meta-analysis of rs-fMRI studies on epilepsy, this study demonstrated, as expected, decreased spontaneous activity in the DMN, including the PCC/precuneus, mPFC, and bilateral IPL. Moreover, we found convergent increased local activity in the right thalamus as well as the MTL and sensorimotor cortex. These results not only provide significant insights into the pathogenesis and mechanism of epilepsy, but also provide potential targets for future precise treatment of epilepsy. Specifically, the use of non-invasive brain stimulation in clinical treatment such as TMS.

### 4.1 Increased activity in the thalamus

Our results demonstrated several regions showing significantly increased activity, including the thalamus, hippocampus, and motor area. Among them, the thalamus is the only region showing spatially overlapped increased activity in both TLE and non-TLE patients by both ReHo and ALFF methods.

The thalamus is a powerful gateway and transmission core that transfers massive sensory information to the cerebral cortex. The thalamus is connected to the cortex via thalamocortical and corticothalamic pathways, which form extensive reciprocal connections (22,23). These connections allow for the bidirectional flow of information between the thalamus and various cortical regions, including the SOZ. It has been suggested that the interaction between the SOZ and the thalamus could contribute to seizure initiation, maintenance, and termination through the modulation of cortical excitability and synchronization (24,25,37,38).

According to the above mechanism, several deep brain stimulation (DBS) studies had taken the thalamus as a target and reported a significant control of the seizures in drug-resistant epilepsy patients (39,40). Specifically, systematic review and meta-analysis data has suggested better effects of DBS in the anterior thalamic nucleus (ANT) for focal seizures, while a better effect in the centromedian thalamic nucleus (CMT) for generalized seizures (41,42). These DBS results inspired our hypothesis of increased activity in the thalamus. We found that the peak point was in mediodorsal lateral thalamus, between ANT and CMT (SuppFig 3). Our results do not seem to be precisely consistent with target selection for DBS. In fact, there has been lack of direct evidence for the selection of DBS target for the treatment of drug-resistant epilepsy.

Thus, we were interested in the original studies that reported increased local activity in the ANT or CMT. Among the 8 studies reporting increased ReHo or ALFF in the thalamus, only one study reported peak activity in the ANT, and no study reported peak activity in the CMT (SuppTab 1). Most of the reported clusters were located in mediodorsal thalamus. However, considering that the spatial resolution of fMRI is limited (usually more than 3 mm during scanning), and all studies performed spatial smoothing for data analysis, the spatial location of the current meta-analytic result may not be accurate enough for guiding the target selection of focused therapy. Moreover, the spatial distance among the three thalamus nuclei is actually very close. In future studies, using high spatial resolution images and non-smooth analysis for direct comparison are expected to provide more accurate localization.

### 4.2 Decreased activity in the default mode network (DMN)

One of the most prominent features in cognitive processing is task-independent deactivation of the DMN (43,44). It means that during a variety of cognitive tasks, the task related regions show activation, while the DMN regions show task-independent deactivation (43). For example, previous studies have demonstrated that patients with mTLE showed reduced DMN connectivity and memory decline (45), and generalized epilepsy patients showed impaired language function with reduced suppression of DMN (46). Moreover, it had been reported in a review (47) and a meta-analysis (16) that decreased FC in DMN was observed across different epilepsies, which may reflect the inhibition of DMN. It seems that, in epilepsy, similar to the task negative activation, the activity of DMN is reduced, and this reduction may have some impact on cognitive functions.

Given decreased FC by itself does not indicate which specific brain region is abnormal, we focus on the local activity by combining studies with ALFF, ReHo, and ICA-DMN in this meta-analysis. Our finding, to some extent, integrated different types of local activity results, and demonstrated convergent regions of DMN. These results may help us to understand the cognitive impairment due to epilepsy and its pathological mechanism.

### 4.3 Meta-correlation between thalamic activity and DMN activity

This study has revealed, for the first time, a negative correlation between local thalamic activity and default mode network (DMN) activity in patients with epilepsy. While decreased DMN activity has been well-documented in epilepsy studies, it remains unclear which regions are antagonistic to this decrease. Our meta-correlation analysis suggests that the aberrant increase in thalamic activity may play a crucial role in the reduced activity of DMN. Specifically, heightened local thalamic activity is associated with weakened DMN function. These findings support the potential use of targeting the thalamus as a therapeutic strategy for individuals with epilepsy.

### 4.4 TLE and non-TLE patients

Temporal lobe epilepsy is the most common type of focal epilepsy, and nearly half of the recruited studies in this meta-analysis were related to temporal lobe epilepsy. At the same time, our results also showed that many of the brain regions with significantly increased activity were near the temporal lobe. Therefore, the comparison between TLE and non-TLE is particularly meaningful.

As expected, and consistent with our main results, DMN activity declined similarly in both groups. Also, when non-TLE patients were excluded, TLE patients showed more abnormal areas with increased activity in the MTL, but interestingly, non-TLE patients still showed increased activity in the MTL. In addition, there were areas of abnormal increases in the thalamus that overlapped spatially in both types, however, there were more increases in the non-TLE group. Overall, the patterns were similar for both groups. The results lead to reflection, perhaps creating subcategorization of different types of epilepsy could be beneficial for the clinical diagnosis of epilepsy. Furthermore, whether there are neural mechanisms that can be similar, even if the types are different.

### 4.5 Limitations

Several limitations should be acknowledged while interpreting our findings. Firstly, we examined local brain activity affected by epilepsy and only recruited relevant rs-fMRI studies, however, the changes of DMN in epilepsy may be caused by affecting cognitive function, which needs further sorted out and explored. Secondly, due to the variety and complexity of epilepsy, this study only carries out a relatively simple classification of TLE and non-TLE studies and general analysis over all the epileptic related changes, more specifics need to be further explored.

## Data Availability

All data produced in the present study are available upon reasonable request to the authors

## Acknowledgements

This work was supported by the National Natural Science Foundation of China (NSFC): 81520108016, 82071537 and Key Medical Discipline of Hangzhou.

## Author Contributions

Yang Qiao: literature search, figures, data collection, data analysis, data interpretation, writing

Cong Fu: literature search, data collection, data interpretation, writing

Na Zhao: literature search, data analysis, data interpretation, writing

Matthew Lock: data interpretation, writing

Zhen Yuan: study design, data interpretation, writing

Yu-Feng Zang: study design, data analysis, data interpretation, writing

