## Supplement figures and tables for "Resting state fMRI reveals pervasive thalamic hyperactivity and default mode network hypoactivity in epilepsy: Systematic review and meta-analysis"

**Supplementary materials**

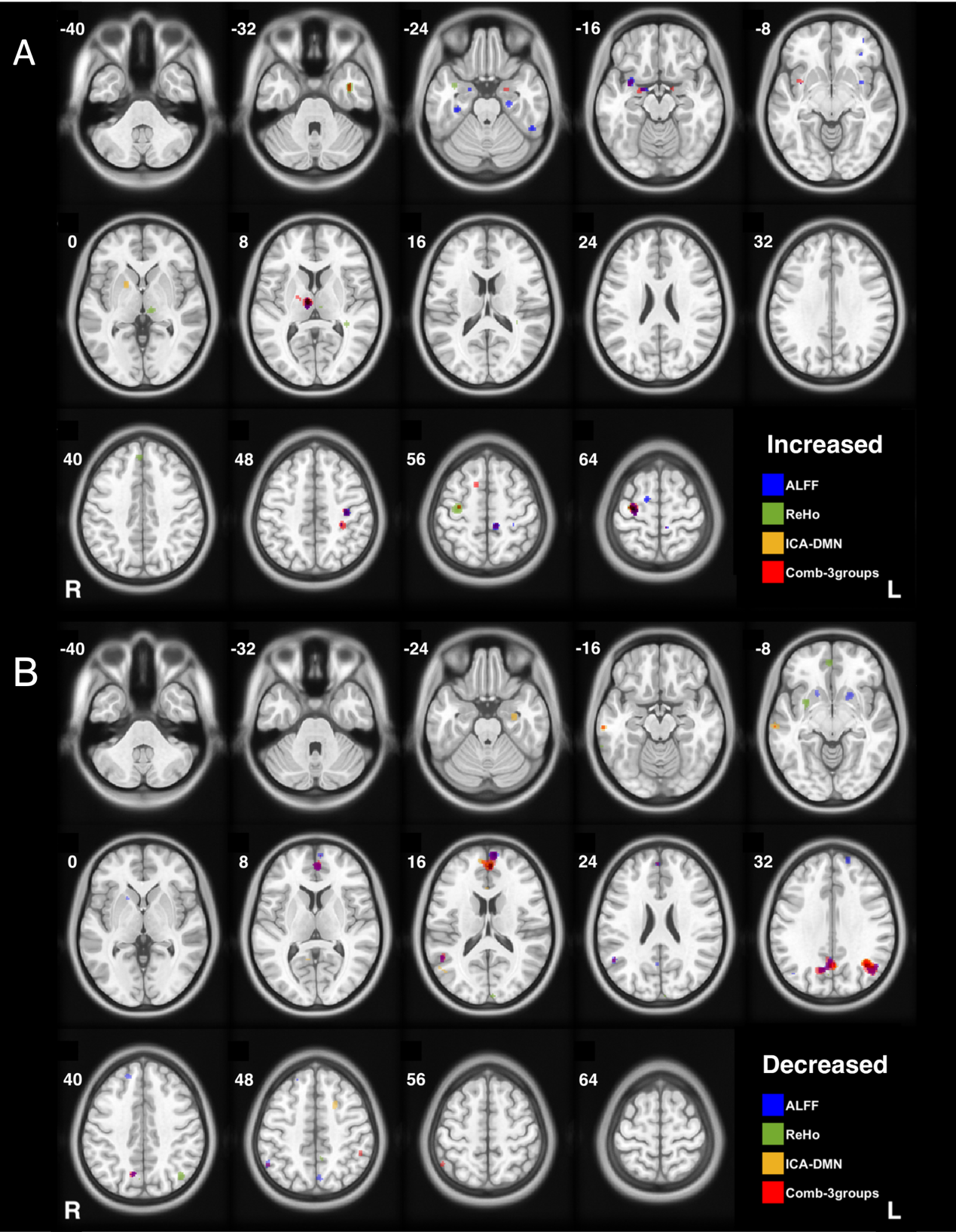

SuppFig 1. ALFF, ReHo and ICA-DMN subgroups meta results compared with the combined meta results. A: regions with increased local activity; B: regions with decreased local activity. Overlay color represented different groups’ results. Blue color indicates ALFF results, green indicates ReHo results, yellow indicates ICA-DMN results and red indicates combined results. The numbers beside each slice represents z coordinate in MNI space. The letters “L” and “R” means left and right separately. “Comb-3groups” indicates results combined all 3 subgroups.

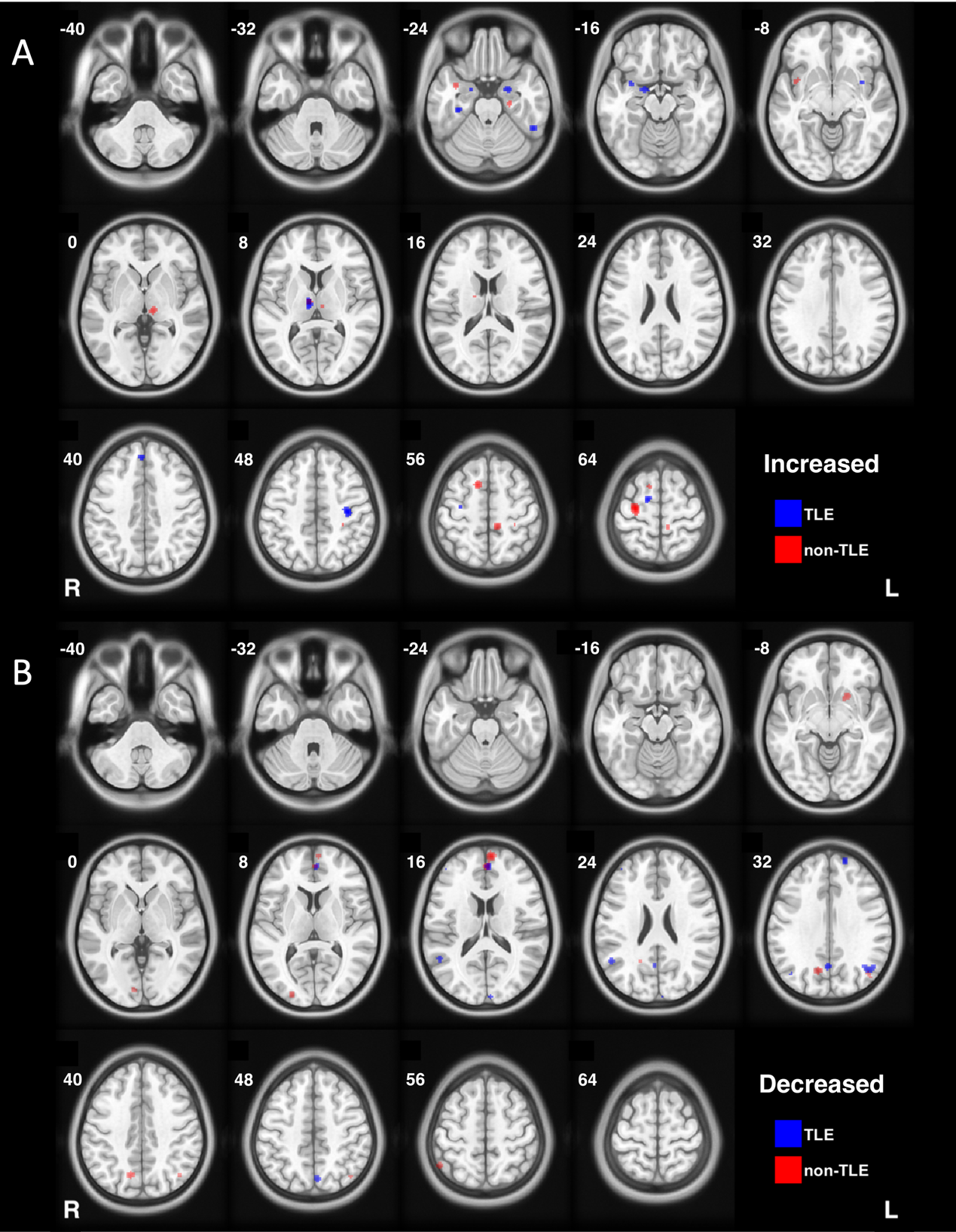

SuppFig 2. TLE and non-TLE groups meta results. A: regions with increased local activity; B: regions with decreased local activity. Overlay color represented different groups’ results. Blue color indicates TLE group results and red color indicates non-TLE group results. The numbers beside each slice represents z coordinate in MNI space. The letters “L” and “R” means left and right separately.

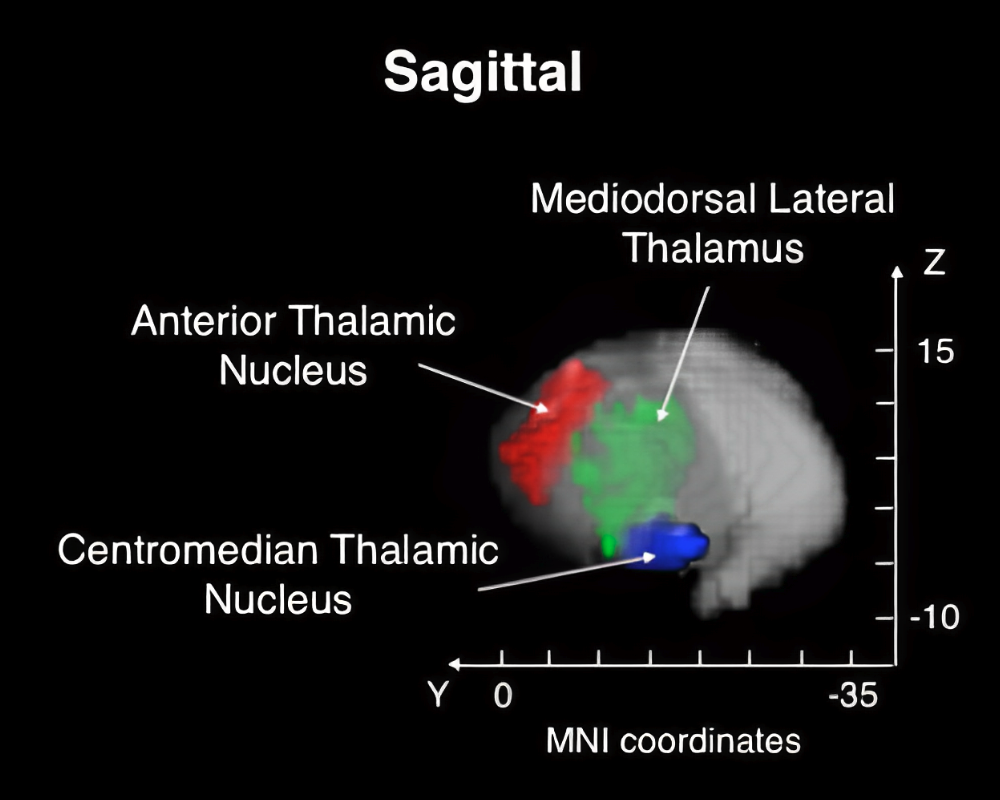

SuppFig 3. Schematic diagram of different thalamic nuclei diagram. Red: anterior thalamic nucleus (highly encouraging DBS location: x [3.8; 5.85], y [−2.1; −6.35] and z [6.2; 10.1] in MNI space). Green: mediodorsal lateral thalamus (peak MNI coordinates of the current meta-analysis results: [8, -12, 8]). Blue: centromedian thalamic nucleus (highly encouraging DBS location: x [5.45±1.15], y [-15.81±1.05], z [-3.25±0.5]).

| Study | Epilepsy type | Cluster Size (mm^3^) | Side | Thalamic subregion | Peak MNI coordinates | | | Cohen's *d* |
| --- | --- | --- | --- | --- | --- | --- | --- | --- |
|  |  |  |  |  | x | y | z |  |
| Zhang 2010 | Mesial temporal lobe epilepsy | 405 | Left | AV | -6 | -3 | 3 | 0.87 |
|  |  | 378 | Right | MDm | 6 | -15 | 6 | 0.65 |
| Zhong 2011 | Generalized tonic-clonic seizures | 1755 | Left | IL | -12 | -21 | 0 | 0.82 |
|  |  | 1188 | right | VL | 9 | -12 | 12 | 1.27 |
| Zhang 2015 | Mesial temporal lobe epilepsy | - | Right | VL | 16 | -9 | 9 | 1.34 |
|  |  | - | Right | VL | 9 | -9 | 9 | 0.97 |
|  |  | - | Left | VL | -15 | -12 | 18 | 1.13 |
|  |  | - | Right | MDl | 9 | -18 | 9 | 0.81 |
| Jiang 2016 | Juvenile myoclonic epilepsy | 1296 | Right | IL | 9 | -21 | 0 | 1.61 |
|  |  | 1026 | Left | MDm | -8 | -23 | 1 | 1.56 |
| Tan 2016 | Infantile spasm | 729 | Right | VL | 15 | -6 | 15 | 1.61 |
|  |  | 729 | Left | VA | -9 | 0 | 9 | 1.79 |
| Qiao 2017 | Idiopathic epilepsy | 756 | Right | PuM | 14 | -30 | 10 | 1.13 |
|  |  | 945 | Left | PuM | -12 | -28 | 9 | 1.38 |
| Wang 2018 | Generalized tonic-clonic seizures | 1350 | Right | VPL | 21 | -18 | 6 | 1.28 |
|  |  | 216 | Left | VL | -24 | -15 | 0 | 1.55 |
|  |  | 216 | Left | VL | -15 | -12 | 3 | 1.49 |
| Liu 2019 | Generalized tonic-clonic seizures | 1863 | Right | MDl | -7 | -17 | 8 | 0.73 |
|  |  | 864 | Left | MDl | 6 | -12 | 8 | 0.59 |

SuppTab 1. Peak thalamus coordinates for studies reporting increased ReHo or ALFF in the thalamus. All thalamic subregions were determined according to the AAL3 template. AV: Anteroventral; IL: Intralaminar; MDm: Mediodorsal Lateral; MDl: Mediodorsal Lateral; PuM: Pulvinar Medial; VA: Ventral Anterior; VL: Ventral Lateral; VPL: Ventral Posterolateral; MNI: Montreal Neurological Institute.
